# A cross-sectional study of Q fever in Camels: risk factors for infection, the role of small ruminants and public health implications for desert-dwelling pastoral communities

**DOI:** 10.1101/2022.04.27.22274356

**Authors:** Peter Holloway, Matthew Gibson, Stephen Nash, Tanja Holloway, Jacqueline Cardwell, Bilal Al Omari, Ehab Abu-Basha, Punam Mangtani, Javier Guitian

## Abstract

Q fever represents an important ‘neglected zoonosis’, with high prevalences recorded across the Middle East region. Among rural desert-dwelling communities in the region, camel milk is largely consumed raw, due to perceptions of dromedaries as a uniquely clean livestock species mentioned in the Qur’an and Islamic hadith, while milk from other livestock species is usually boiled. As a result, camels present a unique public health threat among such communities from milk-borne pathogens, including *C. burnetii*. In view of this, an epidemiological survey was conducted among dromedary herds in southern Jordan between September 2017 and October 2018, including 404 camels from 121 randomly selected herds. In addition, 510 household members associated with these herds were interviewed regarding potential high-risk practices for zoonotic transmission. Weight adjusted camel population seroprevalence for *C. burnetii* was 49.6% (95% CI: 44.7 – 54.5), with evidence of maternally derived immunity in calves ≤6 months old. Adjusted herd-level prevalence was 76.0% (95%CI 72.7–80.2), with 30.4% (144/477) of individuals estimated to consume raw milk from infected herds monthly or more. Following multivariable logistic regression analysis, seropositive status in camels was found to be associated with increasing age, high herd tick burdens, keeping the herd together throughout the year including when calving, and owning larger (>50) sheep and goat flocks, with goats presenting a higher risk than sheep. Racing camel status was found to be protective. Socioculturally appropriate interventions aimed at raising awareness of potential risks associated with drinking raw camel milk, alongside appropriate livestock management interventions, should be considered.

## Introduction

Q fever represents an important ‘neglected zoonosis’, which despite the presence of licensed vaccines, remains largely unrecognized and uncontrolled, particularly among lower and middle-income countries (LMIC) where seroprevalences are often high (*1*). The causative agent, *Coxiella burnetii*, is an obligate gram-negative intracellular bacterium of high tenacity, favouring hot dry conditions, with high infectivity (*2*). Human infections range from being asymptomatic to causing an acute non-specific febrile illness, often with hepatitis and atypical pneumonia (*3*). While most clinical infections are self-limiting, some individuals go on to develop chronic disease, which may include endocarditis and fatigue (*4,5*). These non-specific and diverse signs and symptoms, compounded by a lack of awareness among many healthcare workers and lack of routine laboratory testing in many LMIC settings, mean that individuals presenting with clinical *Coxiella burnetii* infection are frequently misdiagnosed (*6,7*).

In ruminants, Q fever is an important production disease causing reproductive losses through abortions, stillbirths and infertility, alongside milk drop and chronic mastitis (*8*). Bacteria are shed in high numbers through infected birth products, as well as in milk, faeces and urine (*9*). Livestock and human infections occur via inhalation of contaminated dust particles, including infected tick faeces, as well as through contact with infected birthing products and from infected tick bites (*10*). Zoonotic transmission also occurs via consumption of infected raw dairy products (*11*). While the zoonotic impact of *C. burnetii* infection in small ruminant and cattle populations has been widely reported, the potential role of camels in zoonotic transmission of Q fever remains largely unexamined, particularly in the Middle East region, where favourable conditions for the pathogen exist (*12*). The widespread consumption of raw camel milk across the Arab world, due to the perceptions of camels as uniquely clean livestock with mention in the Qur’an and Islamic hadith, means that camels present a unique public health threat (*13,14,15*).

To improve understanding of the epidemiology and potential zoonotic risks posed by Q fever in camels, we conducted a large-scale epidemiological survey among camel herds in southern Jordan, largely owned by desert-dwelling Bedouin communities. This population is considered likely to be representative of analogous Bedouin and pastoral communities in the wider region. The objectives of the study were to: i) estimate the prevalence of *C. burnetii* in the camel population in southern Jordan ii) identify potential transmission pathways for *C. burnetii* infection in camels, particularly regarding the role of small ruminants, and iii) assess the potential public health risk associated with these herds through consumption of raw milk and other activities.

## Methods

### Study design and study population

A cross-sectional study was conducted between 28th October 2017 and 11th October to 2018, in Aqaba and Ma’an governorates of southern Jordan, an area of approximately 40,000 km^2^ and 8,000 camels (based on MoA data) (figure 1). Probabilistic sampling was conducted using camel owner lists supplied by the Ministry of Agriculture (MoA) according to four local administrative areas (Aqaba east, Aqaba west, Ma’an east and Ma’an west). To facilitate owner compliance no more than 12 camels were sampled per herd and in herds of less than 12 all camels were sampled, subject to accessibility and owner permissions. Two standardised structured questionnaires regarding potential risk factors for *C. burnetii* infection, in camels and humans respectively, were administered in the local dialect on Android tablets, using the application Open Data Kit (ODK), among herd owners and their household members. All camels included in the study were clinically examined by a veterinary surgeon to assess general health and the presence of ticks (yes/no), prior to collection of a serum sample.

**Figure 1.**
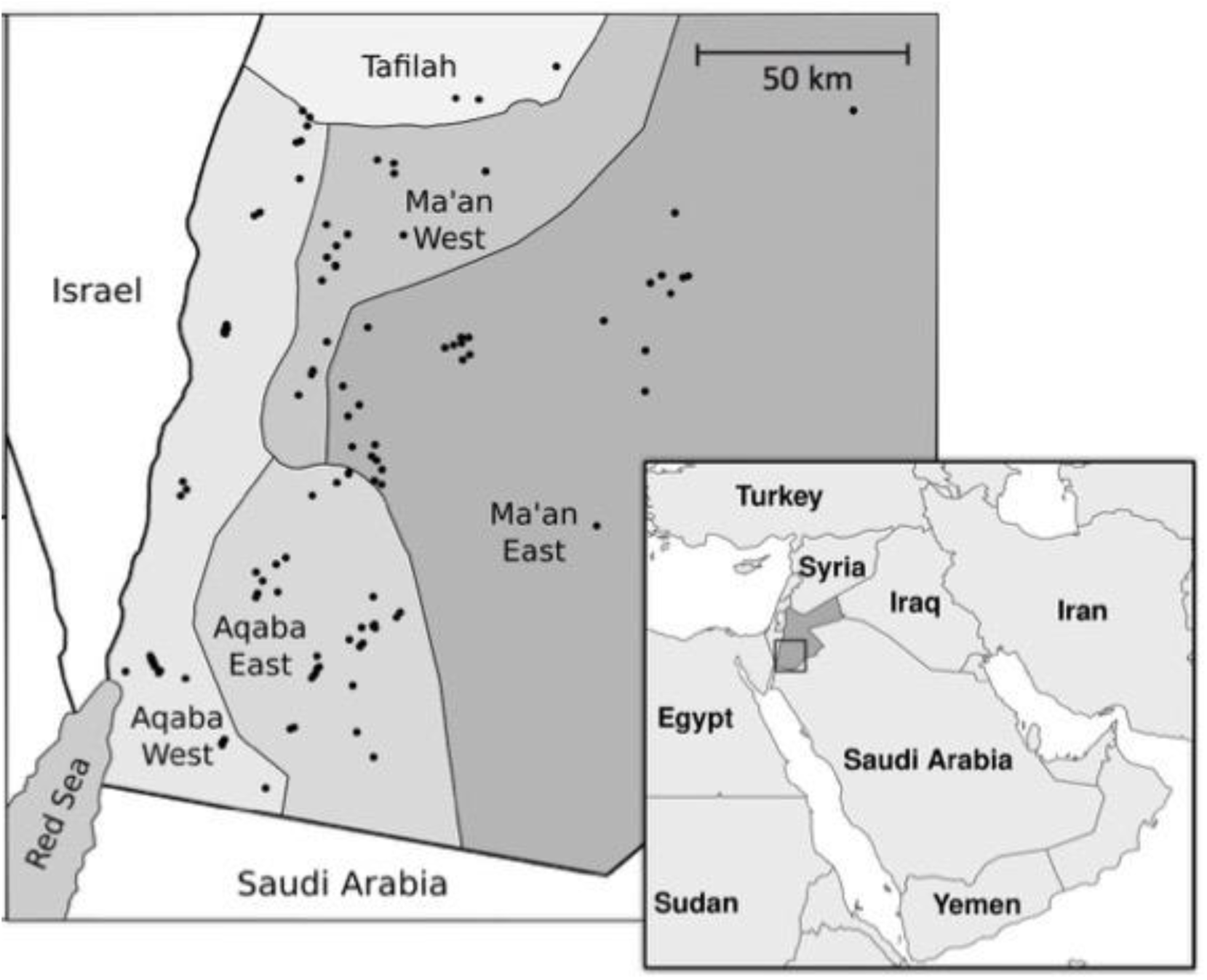
Location of 121 camel herds sampled in southern Jordan October 2017 to October 2018 (due to local grazing movements there were three herds selected from the MoA list for Ma’an west that were sampled in the neighboring region, Tafilah. Results from these herds were attributed to Ma’an west).

### Laboratory methods

Blood samples were collected in 8ml serum vacutainer tubes, transported in cool boxes and centrifuged at 2000 RPM for 10 minutes, followed by serum collection and storage at -20°C. Laboratory testing was performed at the Diagnostic Laboratory, Veterinary Health Centre, Jordan University of Science and Technology, Ar-Ramtha, Irbid, using an indirect ELISA (ID Screen Q fever Indirect Multi-species, IDVet, Montpellier, France). Plates were read at 405 nm and samples with an optical density (OD) of ≥50% were considered positive, according to the manufacturer’s recommendations. Sensitivity and specificity were reported by the manufacturer as being approximately 100%; however, although widely used in camelids, the test has only been validated for use in sheep, goats and cattle (*16*).

### Statistical analysis

We calculated seroprevalence estimates, weighted according to sample size, relative to the estimated camel population, based on MoA data for each sub-region. Regression models were built for identification of risk factors, with camels ≤ 6 months of age excluded from analyses due to the potential influence of maternally derived antibodies.

Univariable analyses were conducted, using mixed-effects logistic regression to adjust for herd-level random effects, with camel serological status considered a binary outcome. All potential risk factors were analysed as categorical variables, with the exception of camel age, altitude of the holding, and small ruminant flock size which were analysed as continuous variables. Season was not considered for analysis due to the non-longitudinal nature of the study and likely correlation with sample location. A composite variable ‘closed herd’ was constructed, defined by herd owners answering ‘no’ to borrowing, lending, purchasing, racing and contact with local or distant herds.

Variables associated with the outcome with a p-value less than 0.2 were considered for inclusion in the multivariable models, with the exception of any variables missing more than 10% of their values. Collinearities between variables were examined using the Pearson R coefficient and a threshold of 0.4, with collinear variables excluded from the same multivariable model. Multivariable models were constructed using a backwards stepwise method, with the least significant variable removed at each step while p > 0.1, unless the variable was considered an a priori factor (sex and age) or the removal of the variable demonstrated a significant effect on the other variables (a change in log odds > 20%). Model building was repeated using a forwards stepwise method, beginning with a priori variables and adding new variables in order of significance, keeping variables if they showed p <0.1 or changed the log odds of other risk factors by > 20%.

The herd-level prevalence of *C. burnetii* (the proportion of herds with at least one camel with antibodies against *C. burnetii* in the serum) was estimated taking into account the uncertainty arising from sampling of only a proportion of each herd (the proportion being different in each herd). The following method was used, based on that described by Beauvais et al. (*17*):

#### Step 1

For each herd, a probability distribution *P*_*i*_, *i=0*…*m* of the number of positive animals (animals with serum antibodies) for herd size *m* was obtained as:

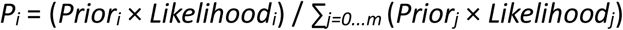

where *Prior*_*i*_ is the prior probability of having i positives in the herd, and *Likelihood*_*i*_ is the likelihood of having i positives given the sample results for that herd. Then, the probability of the herd having at least one positive *P*_*+*_ is 1-*P*_*0*_. The prior distribution for the number of positives in the herd was calculated separately for each herd, as described by Beauvais et al (19). A frequency distribution of within-herd prevalences was multiplied by the number of camels in the particular herd, and the results were rounded to whole numbers to give a discrete probability distribution for positives within the herd.

#### Step 2

Each herd was simulated as being positive or negative using a random sample from a binomial distribution with probability of success *P*_*+*_. This was repeated 10000 times to create an uncertainty distribution, where the 2.5th and 97.5th percentiles give a 95% credible interval and the 50th percentile gives the most likely herd-level prevalence.

The number of individuals living in households with Q fever positive herds was compared against questionnaire data relating to potential pathways for *C. burnetii* zoonotic transmission, by calling a herd positive when there was at least a 50% probability (with 95% confidence) of the herd having at least one positive animal, using Bayesian probability. This figure was used to a calculate the percentage of the sample population likely to have been exposed to potential *C. burnetii* transmission via high-risk practices.

All statistical analyses were performed in R (version 3.5.6) with mixed-effects models generated using the glmer function of the package lme4 (version 1.1-23).

### Informed consent

Informed consent was obtained from all participating camel owners and household members at the time of sampling. Institutional and national guidelines for care, use, and handling of animals were followed at all times. Studies were conducted with institutional ethical review board approval by the Royal Veterinary College, London School of Hygiene and Tropical Medicine (London, UK) and Jordan University of Science and Technology (Irbid, Jordan).

## Results

Blood samples were collected from 404 camels in 121 herds, with an average of 3.3 camels sampled per herd (median herd size 9, IQR 4 - 17). The questionnaire regarding potential risk factors for infection in camels was administered to all 121 herd owners, while the questionnaire regarding potential high-risk practices for human infection was administered to 510 members of camel-owning households (which included the 121 herd owners). Camel numbers sampled were: Ma’an east 90 (29 herds), Ma’an west 69 (21 herds), Aqaba east 147 (36 herds) and Aqaba west 90 (35 herds). MoA records described an estimated 1909 camels (138 herds) in Ma’an East, 1405 camels (127 herds) in Ma’an West, 3563 camels (198 herds) Aqaba East and 873 camels (119 herds) in Aqaba West. Model outcomes were thus weighted for each region by 2.02, 0.65,0.66, 0.80, respectively.

Of the 404 camels sampled there were 8 samples with insufficient serum. Of the remaining 396 samples, 189 were seropositive for *C. burnetii*, giving an unadjusted seroprevalence of 47.7% and a weighted seroprevalence of 49.6% (95% CI: 44.7 – 54.5). Of these, 39 camels were aged ≤ 6 months with 18 seropositive (46.2%), OR 5.1; 95% CI 2.1–12.8; p < 0.01 compared with camels >6 months –2 years of age of whom 14.5% (11 positive / 76 total) were seropositive, with seroprevalence then increasing with age (figure 2). Following removal of calves ≤ 6 months old from the data set, the adjusted seroprevalence was 49.3% (95% CI: 44.0 – 54.6) among 119 herds. Ticks were observed on 226 (55.9%) of camels sampled.

**Figure 2.**
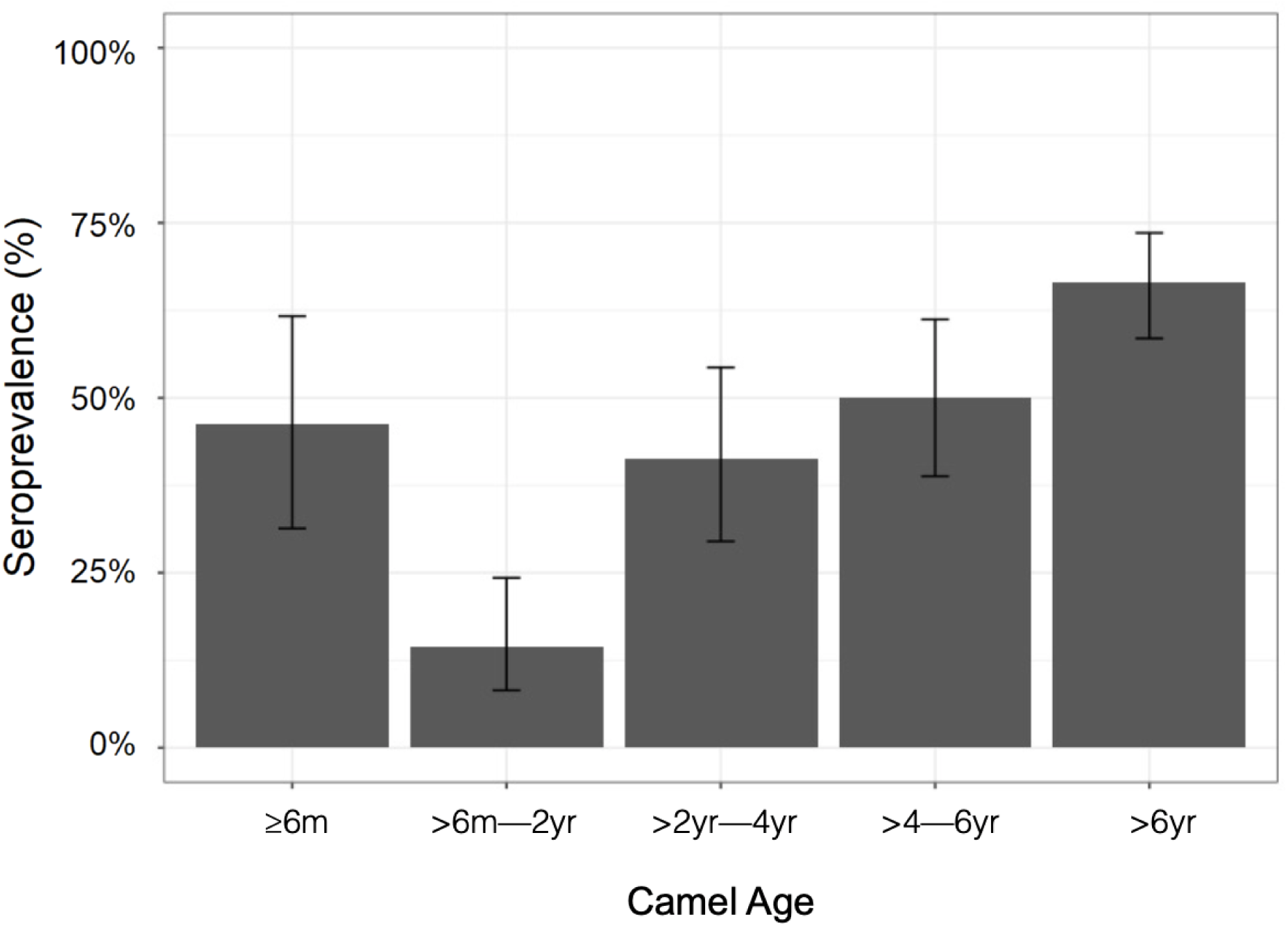
C. burnetii seroprevalence among camel populations in southern Jordan, October 2017 to October 2018, stratified by age.

Descriptive statistics and univariable model results are presented in table 1 and figure 2. Significant correlations, where R>0.4, were identified between the variables ‘region’ & ‘altitude’, between ‘herd owner has small ruminants (linear per 10)’ and ‘herd owner has >50 sheep’ and ‘herd owner has >50 goats’, and between ‘closed herd’ and ‘contact with local camels’ (inverse).

**Table 1.**
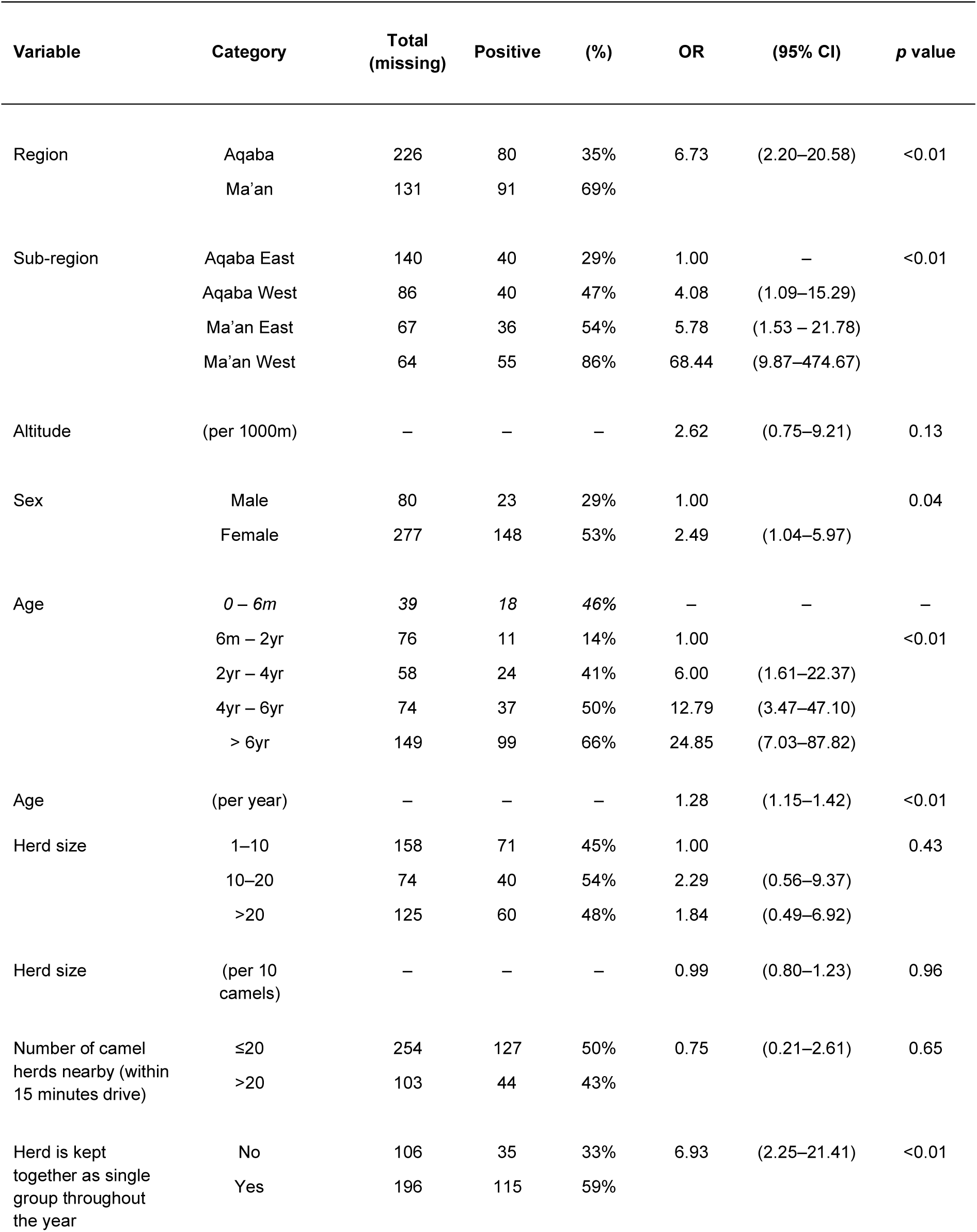
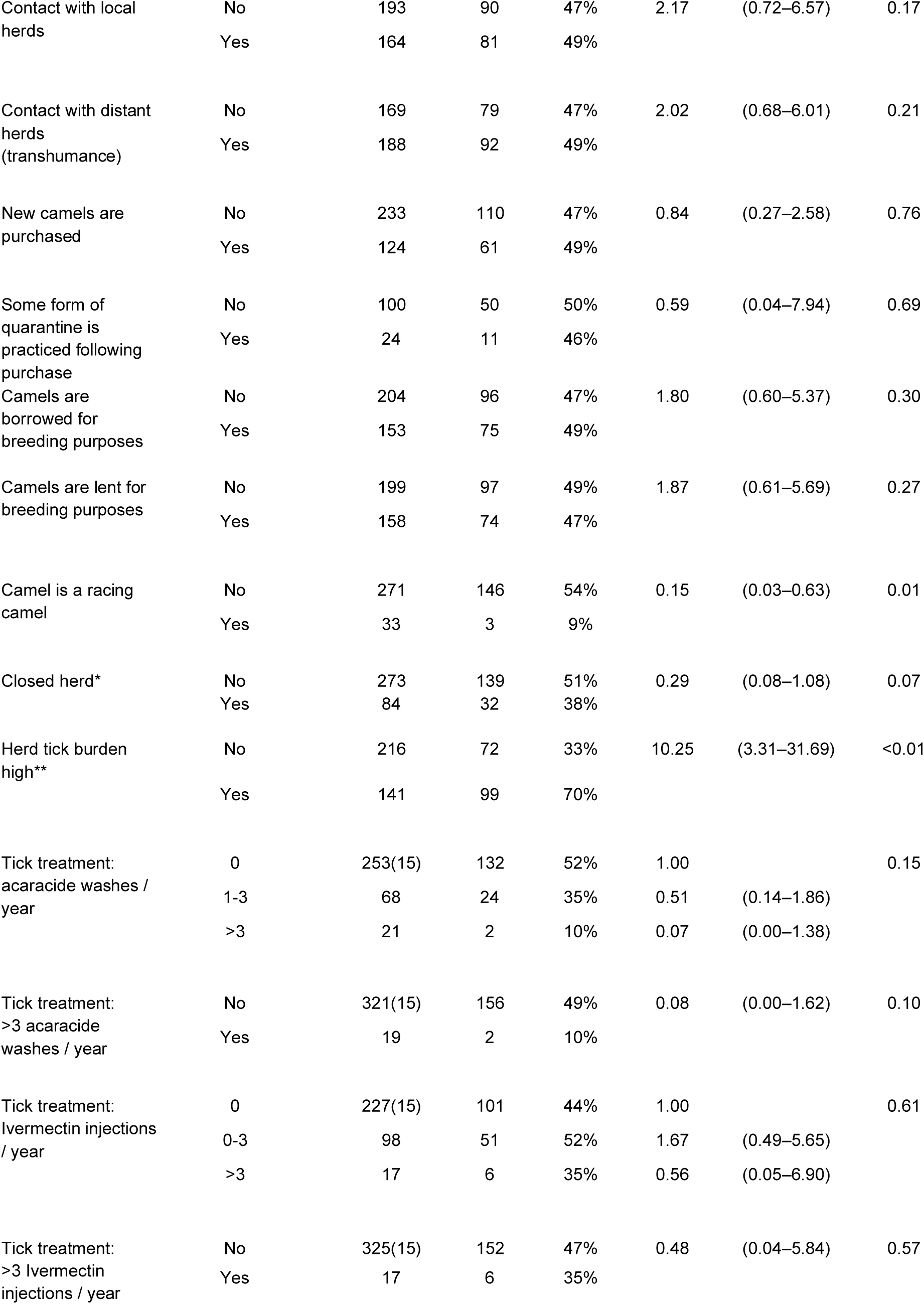
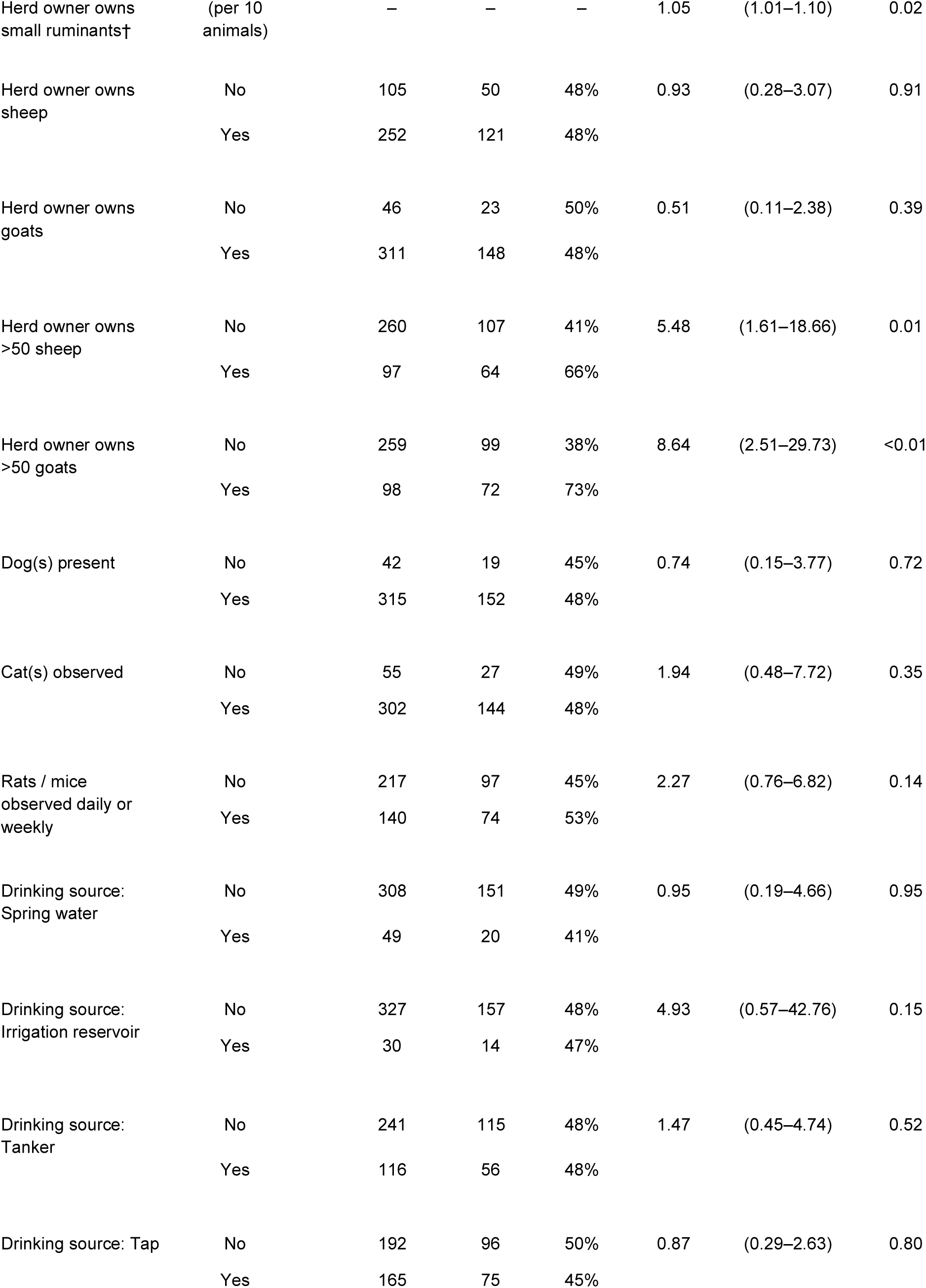
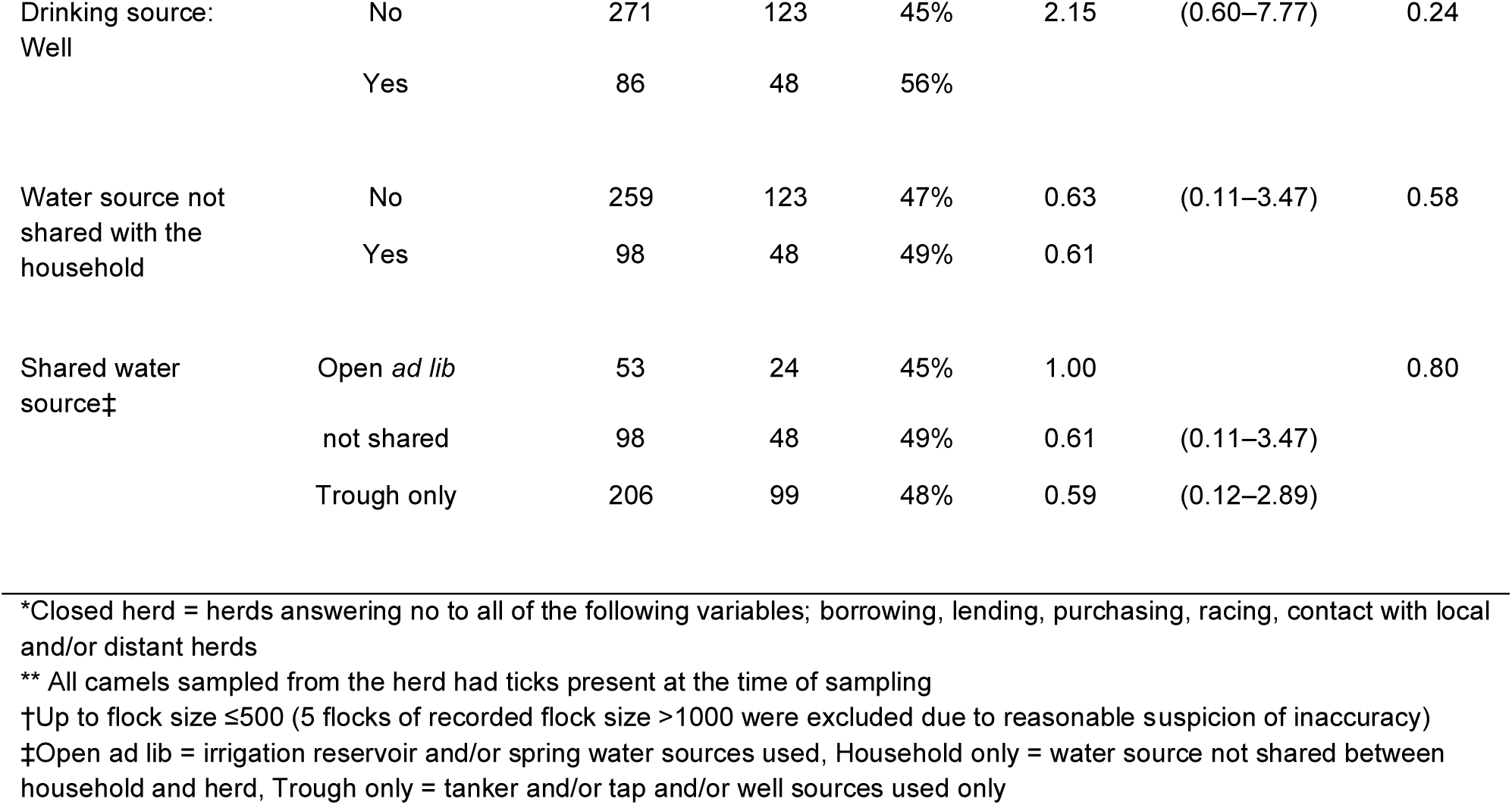
Characteristics of study sample and univariate associations between potential risk factors and C. burnetii seropositivity in camel populations in southern Jordan, October 2017 to October 2018 (due to the potential influence of maternal immunity camels ≤6m have been excluded from all variables except age).

Multivariable model results are shown in Table 2, with the variable, ‘camel is a racing camel’ found to be protective (OR_adj_ 0.14; 95% CI 0.03–0.62; *p*=0.012). There was evidence of positive association between *C. burnetii* seropositivity and increasing age, per year (OR_adj_ 1.26; 95% CI 1.14–1.42; *p*<0.001), high herd tick burden (ORadj 7.90; 95% CI 2.50–19.29; *p*<0.001), herd kept as a single group throughout the year (OR_adj_ 5.93; 95% CI 1.52–10.77; *p*=0.006). While simply owning sheep and goats was not significantly (<0.05) associated with risk, increasing flock size (linear, per 10) (OR_adj_ 1.10; 95% CI 1.00–1.09; *p*=0.037), and owning larger flocks were associated: flock size >50 goats (OR_adj_ 8.78; 95% CI 2.04–18.06;) *p*=0.001) and flock size >50 sheep (OR_adj_ 5.22; 95% CI 1.20–11.24; *p*=0.024). Due to significant collinearity (R>0.4), these latter two variables were analysed in separate models, in place of the variable ‘herd owner has small ruminants’ (linear, per 10); all other variables maintained in the final model continued to demonstrate significant association (p<0.05) with *C. burnetii* seropositivity. Also due to collinearity (R>0.4), the variables ‘closed herd’ and ‘contact with local herds’ were included in separate multivariable models, though neither was maintained in the final model.

**Table 2.** Multivariable associations between potential risk factors and C. burnetii seropositivity in camel populations in southern Jordan, October 2017 to October 2018 (due to the potential influence of maternal immunity camels ≤6m have been excluded).

| Variable | Category | A-priori adjusted OR<br>(95% CI)<sup>1</sup> |  | p value | Fully adjusted OR<br>(95% CI)<sup>2</sup> |  | p value |
| --- | --- | --- | --- | --- | --- | --- | --- |
| **Age** | **(per year)** | 1.26 | (1.14–1.40) | <0.001 | **1.26** | **(1.14–1.42)** | **<0.001** |
| **Herd tick burden high\*** | **Yes** | 7.90 | (2.69–23.18) | <0.001 | **7.90** | **(2.50–19.29)** | **<0.001** |
| **Herd kept as single group all year** | **Yes** | 5.93 | (2.06–17.10) | <0.001 | **5.93** | **(1.52–10.77)** | **0.006** |
| **Herd owner also owns small ruminants†** | **(per 10)** | 1.05 | (1.01–1.09) | 0.017 | **1.04** | **(1.00–1.09)** | **0.037** |
| **Camel is a racing camel** | **Yes** | 0.14 | (0.03–0.63) | 0.010 | **0.14** | **(0.03–0.62)** | **0.012** |
| Sex | Female | 1.62 | (0.68–3.86) | 0.281 | 1.62 | (0.36–2.35) | 0.874 |
| Altitude** | (per 1000m) | 1.90 | (0.58–6.21) | 0.286 | – | – | – |
| Contact with local herds | Yes | 1.86 | (0.67–5.20) | 0.237 | – | – | – |
| Closed herd*** | Yes | 0.27 | (0.08–0.93) | 0.039 | – | – | – |
| Tick treatment:<br>>3 acaricide washes/ yr. | Yes | 0.16 | (0.01–2.88) | 0.215 | – | – | – |
| Rats / mice observed<br>daily or weekly | Yes | 2.19 | (0.79–6.07) | 0.131 | – | – | – |
| Drinking source:<br>Irrigation reservoir | Yes | 7.23 | (0.98–53.45) | 0.053 | – | – | – |
| **Herd owner owns >50 goats†** | **Yes** | 8.78 | (2.78–27.77) | 0.010 | **8.78** | **2.04–18.06** | **0.001** |
| **Herd owner owns >50 sheep†** | **Yes** | 5.22 | (1.66–16.38) | 0.005 | **5.22** | **1.20–11.24** | **0.024** |

<sup>1</sup> Adjusted for a-priori variables; age and sex.
\*All camels sampled from the herd had ticks
<sup>2</sup> Adjusted for; Sex, Age, Herd tick burden high, Herd kept as a single group all year, Herd own owns small ruminants, Camel is a racing camel.
\*\*Due to significant collinearity ( $R>0.4$ ) between 'region' and 'altitude', altitude was chosen over region for inclusion in the final model
\*\*\*Closed herd = herds answering no to; 'borrowing, lending, purchasing, racing, contact with local and/or distant herds.' Due to significant collinearity ( $R>0.4$ ), the variables 'closed herd' and 'contact with local herds' were included in separate multivariable models, though neither variable was maintained in the model.
†Due to significant collinearity ( $R>0.4$ ) between the variables 'Herd owners owns small ruminants (linear, per 10)', 'Herd owner owns >50 sheep' and 'Herd owner owns >50 goats', these variables were included in separate multivariable models. In these models, all variables listed continued to demonstrate significant association ( $p<0.05$ ) with *C. burnetii* seropositivity.

At the herd level, 76.0% (95% credible interval 72.7–80.2) were estimated as being positive for Q fever (having at least one *C. burnetii* seropositive camel present in the herd). It was estimated that 30.4% (145/477) of individuals in camel-owning households were frequently (monthly or more often) drinking raw milk from (their own) Q fever positive herds. In addition, in the past year, 18.8% (96/510) of individuals had been involved in calving camels, 16.7% (85/510) in handling birthing products, 16.5% (84/510) in cleaning camel pens, 7.6% (39/510) in slaughtering camels, 2.6% (13/494) in handling raw camel meat and 10.4% (53/510) had been bitten by ticks (from their own *C. burnetii* positive herds) (table 3).

**Table 3.**
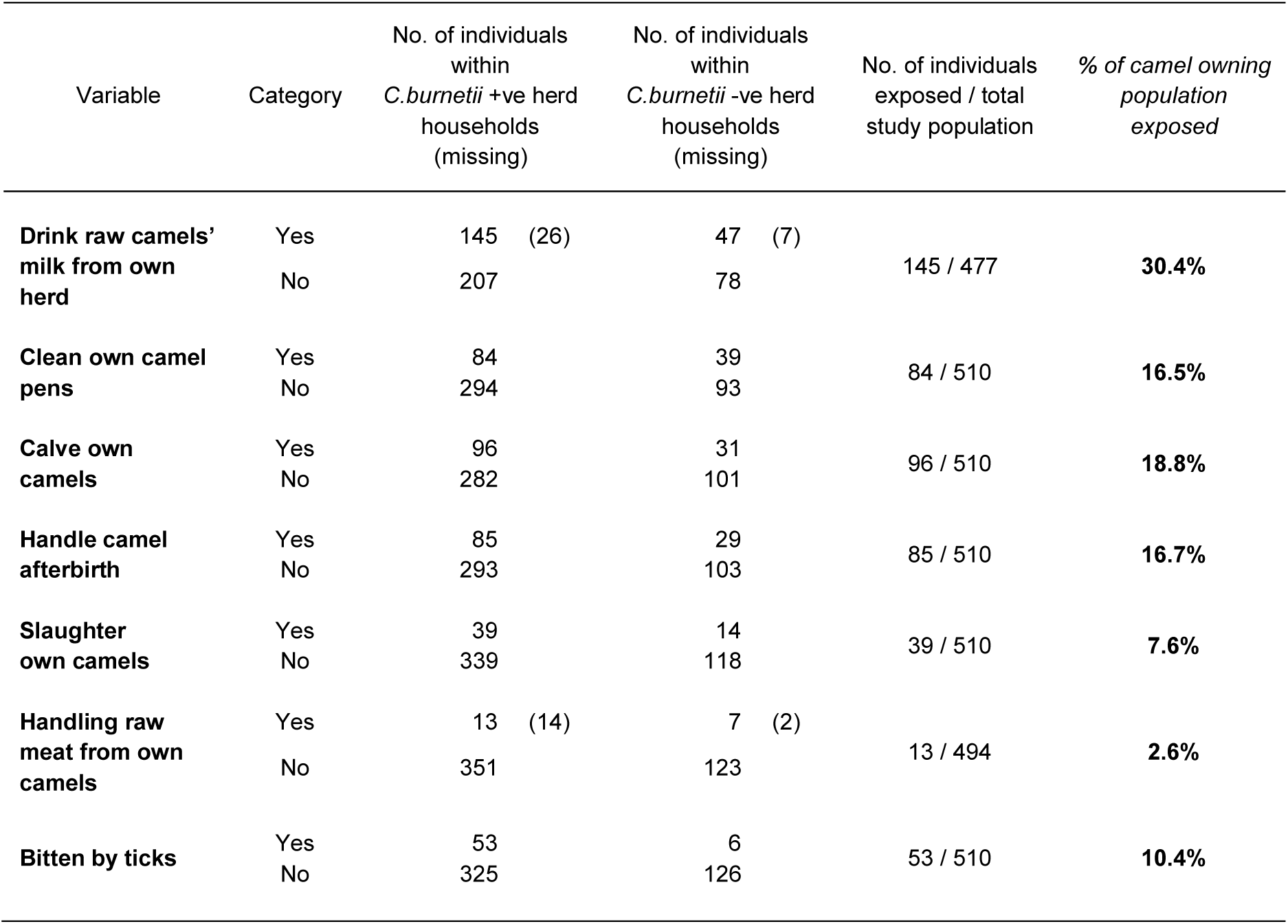
Percentage of study population exposed to potential C. burnetii transmission pathways, using a Bayesian method to predict positive herds with 95% confidence, among camel owning households in southern Jordan, October 2017 to October 2018 (due to the potential influence of maternal immunity, exposure to seropositive camels ≤6m was not included)

| Variable | Category | No. of individuals within<br>*C.burnetii* +ve herd<br>households<br>(missing) | No. of individuals within<br>*C.burnetii* -ve herd<br>households<br>(missing) | No. of individuals<br>exposed / total<br>study population | % of camel owning<br>population<br>exposed |
| --- | --- | --- | --- | --- | --- |
| **Drink raw camels' milk from own herd** | Yes | 145 (26) | 47 (7) | 145 / 477 | **30.4%** |
|  | No | 207 | 78 |  |  |
| **Clean own camel pens** | Yes | 84 | 39 | 84 / 510 | **16.5%** |
|  | No | 294 | 93 |  |  |
| **Calve own camels** | Yes | 96 | 31 | 96 / 510 | **18.8%** |
|  | No | 282 | 101 |  |  |
| **Handle camel afterbirth** | Yes | 85 | 29 | 85 / 510 | **16.7%** |
|  | No | 293 | 103 |  |  |
| **Slaughter own camels** | Yes | 39 | 14 | 39 / 510 | **7.6%** |
|  | No | 339 | 118 |  |  |
| **Handling raw meat from own camels** | Yes | 13 (14) | 7 (2) | 13 / 494 | **2.6%** |
|  | No | 351 | 123 |  |  |
| **Bitten by ticks** | Yes | 53 | 6 | 53 / 510 | **10.4%** |
|  | No | 325 | 126 |  |  |

## Discussion

High seroprevalences of *C. burnetii* are reported in human and livestock populations in the Middle East, where a hot dry climate and open deserts, combined with high, localised small ruminant populations, provide favourable conditions for transmission via dust and wind (*18,19*). However, while the zoonotic risk of Q fever from small ruminants and cattle is well established, the potential risk from camels in the region is poorly understood. The Qur’an describes camels as demonstrating the wisdom and power of God, and the Islamic hadith describe the healing benefits of drinking camels’ milk and urine – meaning that among rural communities in the Arab world, camel milk is usually consumed raw, while the milk from sheep and goats is usually boiled (*15,20*). However, high *C. burnetii* seroprevalences among camel populations in the region (*18*), and detection of *C. burnetii* in raw camel milk samples collected in the region (*21*), suggest consumption of raw camel milk may present an important determinant for Q fever among human populations in the Middle East.

In desert-dwelling pastoral communities, camels and small ruminants are commonly kept together, with potential for pathogen transmission between species (*22*). This means that zoonotic pathogens such as *C. burnetii*, which can be transmitted from small ruminants to camels, present a risk to households owning mixed flocks/herds of small ruminants and camels, via the consumption of raw camel milk, even when milk from other livestock species is boiled. Lafi et al. reported high *C. burnetii* seroprevalences in goats and sheep in Jordan, estimated at 43.3% and 27.0% respectively, and Obaidat et al. have reported a seroprevalence of 24.2% among human populations in Jordan (*18,23*). While Q fever seroprevalences have been reported in camel populations in Tunisia (44.4%), Egypt (21.9%) and Iran (28.7%), our study is the first to report a seroprevalence estimate for camels in Jordan (49.6%; 95% CI: 44.7 – 54.5) (*22, 24,25*).

In the study population, almost a third of camel owners and their households were found to be frequently drinking raw camel milk from *C. burnetii* positive herds. This indicates a clear public health risk from Q fever in camels in the region – alongside other risk-associated camel-engagement activities such as cleaning pens, handling afterbirth, facilitating calving, slaughtering, handling raw meat or camel-tick bites (*26,27*). In addition, high *C. burnetii* seroprevalences in camels suggest likely production losses through infertility, abortions, still-births, weak off-spring, milk drop or chronic mastitis, all with important economic impact (*8*).

Where camels and small ruminants were owned together, simply owning sheep and/or goats was not found to be associated with a significant increase in *C. burnetii* seroprevalence in these camel herds. However, larger small ruminant flock sizes were significantly associated with higher *C. burnetii* seroprevalence in camel herds associated with these flocks. Small ruminant flock sizes of >50 sheep or goats were associated with increased *C. burnetii* seroprevalence in associated herds. In addition, the risk associated with large goat flocks was greater than that associated with large sheep flocks, this being consistent with higher seroprevalences previously identified in goat flocks in the region, and with previous findings identifying goats as posing a greater risk of *C. burnetii* infection than sheep (*18,28*).

Our study findings suggest that controlling Q fever in small ruminant populations is likely to be important in reducing *C. burnetii* seroprevalence in associated camel populations. An effective livestock vaccine for Q fever, Coxevac, manufactured by Ceva, Libourne, France), has been used extensively in the Netherlands (*29,30*). The potential use of such a vaccine among small ruminants and cattle in Jordan and the wider region should be considered. In addition, efficacy studies for possible use of the vaccine in camels should be conducted. This study is the first to demonstrate evidence of maternal antibodies to *C. burnetii* in camels, lasting until approximately 6 months of age. This suggests that, when considering future efficacy studies for existing *C. burnetii* vaccines in camels, vaccination should probably be delayed until after 6 months of age, with further research required. *C. burnetii* seroprevalence increased significantly with camel age, consistent with increased risk of exposure over time and antibodies being long-lasting, in keeping with other studies (*22*).

Due to the high shedding known to occur in small ruminant birthing and aborted materials, small ruminant flocks should be separated from camel herds during the lambing/kidding period where possible (*26*). Indeed, in view of *C. burnetii’s* ability to form tenacious spore-like cell variants, capable of remaining in the environment for more than a year, separate birthing areas for small ruminants and camels are advisable (*8,26*). As the pathogen can travel long distances via the wind, these areas should be as far apart as possible, and moved regularly. The practice of keeping herds together as a single group throughout the year was associated with significantly higher *C. burnetii* seroprevalences in studied herds. This is likely explained by increased exposure to infected camel birthing products in these herds, compared to herds where pregnant females are removed prior to calving (*26*). This suggests removal of pregnant females from the herd prior to calving as a potentially important management intervention in reducing *C. burnetii* transmission between camels. The importance of herd owner hygiene through hand washing (and use of disinfectant foot baths where practical) after working with small ruminants at parturition time, should also be stressed. Such interventions also have relevance in protecting camels (and individuals themselves) from other zoonoses in the region of public health concern, particularly brucellosis, endemic in Jordan and the wider region (*11,31,32*).

Ticks are known to play an important role in *C. burnetii* transmission in livestock populations globally (*26*). In our own study, a high herd tick burden (meaning all camels sampled from the herd had ticks) was significantly associated with higher herd *C. burnetii* seroprevalences. *C. burnetii* has been identified in camel ticks across Africa and the Middle East, particularly in *Hyalomma ssp, Amblyomma spp*. and *Rhipicephalus spp. (33,34*). This suggests that aggressive tick control, using frequent acaracide washes or pour-ons (for example monthly or every two months), could play an important role in controlling Q fever in camel populations in Jordan and the wider region; further research on this is required (*35*). In addition, parallel tick control in small ruminant populations associated with these herds, for example through quarterly dipping, could also be expected to offer a protective effect.

Seroprevalence in racing camels was significantly lower than in non-racing camels, likely explained by separation of racing camels from the main herd and small ruminant flocks, for training purposes. However, parturition during racing lifetime is limited, and racing camels are not usually used in milk production, meaning lower *C. burnetii* seroprevalences are of limited public health impact.

In conclusion, the high seroprevalence of *C. burnetii* in camel herds in southern Jordan coupled with frequent consumption of camel milk and animal husbandry practices where exposure to contaminated environment is high, indicates a clear public health risk. To reduce the zoonotic risk, and to reduce potential production losses, the following targeted management interventions aimed at limiting the transmission of *C. burnetii* in camels should be considered: i) removal of breeding camels from the herd prior to calving, ii) creating separate birthing areas for small ruminants and camels, as far apart as possible and moved regularly, iii) promoting owner biosecurity measures that include hand hygiene (and use of disinfectant foot baths where practical) after working with recently calved or aborted livestock or after handling aborted material. Control measures in small ruminants are of particular importance when managing camel herds alongside larger flocks, particularly regarding goats. Potential licensing and use of existing *C. burnetii* vaccines in Jordan, for use in small ruminants (and cattle), should be considered. In addition, efficacy studies regarding the use of such vaccines in camels should be conducted.

Given the high percentage (over 30%) of individuals in camel-owning households drinking raw camel milk from *C. burnetii* positive herds, educational efforts to promote boiling of camels’ milk should be encouraged. However, in view of the profound cultural barriers likely to be encountered, detailed ethnographic studies to identify public health interventions that are culturally appropriate should first be conducted. In summary, Q fever represents an important zoonosis in the Middle East region and beyond, with high population seroprevalences previously recorded. High *C. burnetii* seroprevalences identified in camels, alongside widespread engagement in high-risk camel-associated practices, including consumption of raw milk, suggest camels likely present a high-risk species for human infection, with culturally appropriate veterinary and public health interventions urgently needed.

## Data Availability

All data produced in the present study are available upon reasonable request to the authors

## Acknowledgements

This research was partly supported by a Medical Research Council Global Challenges Research Fund Foundation award 2017/18 study. The authors would like to sincerely thank the Ministry of Agriculture in Jordan, in particular Dr Fares Ameen Altakhainah, Dr Ghassab Hatem Hasanat, Dr Hassan Hassaen Alhusainat and Dr Abdalmajeed Mahmoud Alajlouni.

## References

1. Vanderburg S, Rubach MP, Halliday JE, Cleaveland S, Reddy EA, Crump JA. 2014. Epidemiology of Coxiella burnetii infection in Africa: a OneHealth systematic review. PLoS neglected tropical diseases 8:e2787.

2. Maurin, M., & Raoult, D. (1999). Q fever. Clinical Microbiology Reviews, 12, 518.

3. van der Hoek W, Versteeg B, Meekelenkamp JC, Renders NH, Leenders AC, et al. (2011) Follow-up of 686 patients with acute Q fever and detection of chronic infection. Clin Infect Dis 52: 1431–1436.

4. Brouqui P, Dupont HT, Drancourt M, Berland Y, Etienne J, et al. (1993) Chronic Q fever. Ninety-two cases from France, including 27 cases without endocarditis. Arch Intern Med 153: 642–648.

5. Ayres JG, Flint N, Smith EG, Tunnicliffe WS, Fletcher TJ, et al. (1998) Post-infection fatigue syndrome following Q fever. QJM 91: 105–123.

6. Buijs SB, Bleeker-Rovers CP, van Roeden SE, Kampschreur LM, Hoepelman AIM, Wever PC, Oosterheert JJ. Still New Chronic Q Fever Cases Diagnosed 8 Years After a Large Q Fever Outbreak. Clin Infect Dis. 2021 Oct 20;73(8):1476–1483.

7. Honarmand H. Q Fever: an old but still a poorly understood disease. Interdiscip Perspect Infect Dis. 2012;2012:131932.

8. Overview of Coxiellosis - Generalized Conditions - MSD Veterinary Manual https://www.msdvetmanual.com/generalized-conditions/coxiellosis/overview-of-coxiellosis

9. Canevari JT, Firestone SM, Vincent G, Campbell A, Tan T, Muleme M, Cameron AWN, Stevenson MA. The prevalence of Coxiella burnetii shedding in dairy goats at the time of parturition in an endemically infected enterprise and associated milk yield losses. BMC Vet Res. 2018 Nov 20;14(1):353.

10. Angelakis E, Raoult D. Q Fever. Vet Microbiol. 2010 Jan 27;140(3-4):297–309.

11. Signs KA, Stobierski MG, Gandhi TN. Q fever cluster among raw milk drinkers in Michigan, 2011. Clin Infect Dis. 2012 Nov 15;55(10):1387–9.

12. Devaux CA, Osman IO, Million M, Raoult D. Coxiella burnetii in Dromedary Camels (Camelus dromedarius): A Possible Threat for Humans and Livestock in North Africa and the Near and Middle East? Front Vet Sci. 2020 Nov 5;7:558481.

13. Qur’an Surra 88, verses 17–20

14. Hadith Sahih Bukhari, Ablutions, Volume 1, Book 4, Number 234)

15. Galali Y, Al-Dmoor H. Miraculous Properties of Camel Milk and Perspective of Modern Science J Fam Med Dis Prev 2019, 5:095

16. ID Screen® Rift Valley Fever Competition Multi-species, https://www.id-vet.com/produit/id-screen-rift-valley-fever-competition-multi-species/

17. Beauvais W, Orynbayev M, & Guitian J. (2016). Empirical Bayes estimation of farm prevalence adjusting for multistage sampling and uncertainty in test performance: a Brucella cross-sectional serostudy in southern Kazakhstan. Epidemiology and infection, 144(16), 3531–3539

18. Lafi SQ, Talafha AQ, Abu-Dalbouh MA, Hailat RS, Khalifeh MS. Seroprevalence and associated risk factors of Coxiella burnetii (Q fever) in goats and sheep in northern Jordan. Trop Anim Health Prod. 2020 Jul;52(4):1553–1559.

19. Ergas D, Keysari A, Edelstein V, Sthoeger ZM. Acute Q fever in Israel: clinical and laboratory study of 100 hospitalized patients. Isr Med Assoc J. 2006 May;8(5):337–41.

20. Abdel Gader A, Alhaider A. The unique medicinal properties of camel products: A review of the scientific evidence. Journal of Taibah University Medical Sciences (2016) 11(2), 98e103

21. Esmaeili S, Mohabati Mobarez A, Khalili M, Mostafavi E, Moradnejad P. Molecular prevalence of Coxiella burnetii in milk in Iran: a systematic review and meta-analysis. Trop Anim Health Prod. 2019 Jul;51(6):1345–1355.

22. Selim A, Ali AF. Seroprevalence and risk factors for C. burentii infection in camels in Egypt. Comp Immunol Microbiol Infect Dis. 2020 Feb;68:101402.

23. Obaidat, M. M., Malania, L., Imnadze, P., Roess, A. A., Bani Salman, A. E., & Arner, R. J. (2019). Seroprevalence and Risk Factors for Coxiella burnetii in Jordan. The American journal of tropical medicine and hygiene, 101(1), 40–44.

24. Selmi R, Mamlouk A, Ben Yahia H, Abdelaali H, Ben Said M, Sellami K, Daaloul-Jedidi M, Jemli MH, Messadi L. Coxiella burnetii in Tunisian dromedary camels (Camelus dromedarius): Seroprevalence, associated risk factors and seasonal dynamics. Acta Trop. 2018 Dec;188:234–239.

25. Janati Pirouz H, Mohammadi G, Mehrzad J, Azizzadeh M, Nazem Shirazi MH. Seroepidemiology of Q fever in one-humped camel population in northeast Iran. Trop Anim Health Prod. 2015 Oct;47(7):1293–8.

26. Devaux, C. A., Osman, I. O., Million, M., & Raoult, D. (2020). Coxiella burnetii in Dromedary Camels (Camelus dromedarius): A Possible Threat for Humans and Livestock in North Africa and the Near and Middle East?. Frontiers in veterinary science, 7, 558481.

27. Mohammadpour R, Champour M, Tuteja F, Mostafavi E. Zoonotic implications of camel diseases in Iran. Vet Med Sci. 2020 Aug;6(3):359–381

28. Vellema P, Santman-Berends I, Dijkstra F, van Engelen E, Aalberts M, Ter Bogt-Kappert C, van den Brom R. Dairy Sheep Played a Minor Role in the 2005-2010 Human Q Fever Outbreak in The Netherlands Compared to Dairy Goats. Pathogens. 2021 Dec 3;10(12):1579.

29. Coxevac, inactivated Coxiella burnetii vaccine, European medicines agency, https://www.ema.europa.eu/en/medicines/veterinary/EPAR/coxevac

30. Hogerwerf, L., van den Brom, R., Roest, H. I., Bouma, A., Vellema, P., Pieterse, M., Dercksen, D., & Nielen, M. (2011). Reduction of Coxiella burnetii prevalence by vaccination of goats and sheep, The Netherlands. Emerging infectious diseases, 17(3), 379–386.

31. Musallam II, Abo-Shehada MN, Hegazy YM, Holt HR, Guitian FJ. Systematic review of brucellosis in the Middle East: disease frequency in ruminants and humans and risk factors for human infection. Epidemiol Infect. 2016 Mar;144(4):671–85.

32. Bardenstein, S., Gibbs, R. E., Yagel, Y., Motro, Y., & Moran-Gilad, J. (2021). Brucellosis Outbreak Traced to Commercially Sold Camel Milk through Whole-Genome Sequencing, Israel. Emerging Infectious Diseases, 27(6), 1728–1731.

33. Mumcuoglu KY, Arslan-Akveran G, Aydogdu S, Karasartova D, Kosar N, Gureser AS, Shacham B, Taylan-Ozkan A. Pathogens in ticks collected in Israel: I. Bacteria and protozoa in Hyalomma aegyptium and Hyalomma dromedarii collected from tortoises and camels. Ticks Tick Borne Dis. 2022 Jan;13(1):101866

34. Getange D, Bargul JL, Kanduma E, Collins M, Bodha B, Denge D, Chiuya T, Githaka N, Younan M, Fèvre EM, Bell-Sakyi L, Villinger J. Ticks and Tick-Borne Pathogens Associated with Dromedary Camels (Camelus dromedarius) in Northern Kenya. Microorganisms. 2021 Jun 30;9(7):1414

35. el-Azazy OM. Camel tick (Acari:Ixodidae) control with pour-on application of flumethrin. Vet Parasitol. 1996 Dec 31;67(3-4):281–4.

